# Integrating Tumor and Organoid DNA Methylation Profiles Reveals Robust Predictors of Chemotherapy Response in Rectal Cancer

**DOI:** 10.1101/2025.02.28.25322951

**Authors:** David Lukacsovich, Wini Zambare, Chao Wu, Hanchen Huang, Wei Zhang, Min Jung Kim, Janet Alvarez, Aron Bercz, Philip B. Paty, Paul B. Romesser, Lily Wang, J. Joshua Smith, X. Steven Chen

## Abstract

Rectal cancer patients display heterogeneous responses to neoadjuvant treatment—including the intensive total neoadjuvant therapy (TNT)—and reliable biomarkers are lacking to guide which tumors will benefit most from these regimens. Here, we profiled DNA methylation in tumor tissue and matched patient-derived organoids (PDOs) from 18 rectal cancer cases (50 total samples), leveraging the Illumina MethylationEPIC array and quality control filters that retained 771,964 CpG sites. Analyses used linear models (for tissue-only or PDO-only) and a joint linear mixed-effects approach (accounting for patient-level random effects) to identify significant CpGs associated with log-transformed FOLFOX IC50. We found that PDOs faithfully recapitulate patient-tumor methylation patterns (Spearman’s correlation >0.95 among replicate organoids), and the joint model uncovered 745 CpGs tied to FOLFOX sensitivity, many of which were missed in tissue-only analyses. Differentially methylated regions reinforced that broader epigenetic blocks near TSS or enhancer regions may modulate chemo-resistance, while pathway enrichment pinpointed focal adhesion, ECM–receptor interaction, calcium signaling, and folate metabolism as key processes. A methylation risk score derived from these CpGs significantly predicted progression-free survival in an independent colorectal cancer cohort (p=0.019), outperforming single-sample–based signatures. These findings suggest that combining methylation profiles from both tumors and PDOs can expose robust epigenetic drivers of therapy response, aiding the development of clinically actionable biomarkers for rectal cancer TNT.

## Introduction

Colorectal cancer (CRC) remains a major health challenge, ranking as the third most commonly diagnosed cancer and the second leading cause of cancer-related death worldwide[1, 2]. Rectal cancer, which comprises a significant fraction of these cases, often requires a multimodal treatment approach. In locally advanced rectal cancer, standard care has evolved to include neoadjuvant therapies – traditionally 5-fluorouracil (5-FU)-based chemoradiotherapy (CRT) prior to surgery. More recently, Total Neoadjuvant Therapy (TNT) has been introduced, delivering both systemic chemotherapy and chemoradiation *before* surgical resection [3, 4]. TNT aims to increase pathologic complete response (pCR) and clinical complete response rates, facilitate sphincter preservation, and tackle micrometastatic disease earlier [5–7]. This intensive approach, however, comes with substantial toxicity and risks. Crucially, not all patients benefit equally: pCR is achieved in only ∼15–20% of cases when surgery is mandated, while up to one-third of patients derive minimal or no tumor regression from neoadjuvant treatment [8]. Identifying predictive biomarkers of response to TNT is therefore a pressing need. An accurate pretreatment predictor could spare non-responders the morbidity of futile therapy and help triage responders to organ-preserving strategies or selective omission of surgery [9]. Despite considerable efforts, no clinically validated biomarker for rectal cancer therapy response exists to date. Prior research has explored diverse candidates – from imaging features and circulating markers to tumor gene expression profiles – but with limited success DNA methylation biomarkers have gained significant attention over the past two decades as potential predictors of treatment response in colorectal cancer (CRC), including rectal tumors. Research on locus-specific hypermethylation (e.g., *MLH1*, *MGMT*) and global changes (e.g., LINE-1 hypomethylation) has revealed strong links between epigenetic alterations and both chemotherapy and radiotherapy outcomes, particularly in neoadjuvant settings [10–14]. In rectal cancer, methylation panels such as CIMP and MINT loci, as well as individual genes (e.g., *TFAP2E*, *TIMP3*, *MGMT*), have shown associations with therapeutic responsiveness, although inconsistent validation underscores the need for larger, multi-institutional studies [15–18]. Recent efforts extend to multi-gene predictive models, liquid biopsy approaches, and epigenetic therapy combinations, highlighting methylation’s potential to refine patient stratification for protocols like total neoadjuvant therapy (TNT) [19–22]. Despite hurdles in reproducibility and tumor heterogeneity, this growing body of evidence supports integrating methylation data into modern treatment paradigms, with the ultimate goal of identifying patients most likely to benefit from intensive regimens and sparing others from unnecessary toxicity.

Patient-derived organoids (PDOs) have emerged as a transformative model to address these gaps in rectal cancer research—particularly for biomarker discovery. Unlike standard cell lines, PDOs are generated directly from patient tumor biopsies or resection specimens, maintaining the original tumor’s histopathologic architecture, mutational spectrum, and clonal heterogeneity[23]. Our group and others have shown that rectal cancer PDOs reflect patient-specific treatment responses, achieving a high success rate (>90%) in establishing stable cultures from both untreated and post-therapy samples [24, 25]. This efficient pipeline not only preserves key oncogenic mutations and epigenetic features but also enables ex vivo testing of standard regimens (e.g., 5-FU, oxaliplatin) and combination therapies—crucial for dissecting TNT resistance [24]. Moreover, by coupling PDO-based drug response assays (e.g., IC50 calculations) with multi-omics analyses (including methylation profiling), we can identify tumor-intrinsic factors conferring chemo- or radioresistance. In our recent study, we demonstrated that integrating matched colorectal tumor and organoid gene expression data significantly improves chemotherapy response prediction for colorectal cancer [26]. Critically, the matched patient-to-PDO framework serves as a biologically enriched “filter” for candidate biomarkers: if a putative methylation marker correlates with response in both the patient’s tumor and its corresponding PDO, it is more likely to represent a true driver of therapy outcome rather than a confounding epiphenomenon. This platform thus accelerates preclinical validation of new targets and biomarker panels, laying the groundwork for eventual clinical integration—where validated methylation signatures could guide treatment selection without requiring each patient to undergo a full PDO derivation in the clinic.

Although TNT includes both chemotherapy and radiotherapy, we focus here on chemotherapy response (IC50) as a surrogate for overall TNT efficacy because systemic chemotherapy is central to TNT protocols, and tumors resistant to 5-FU–based regimens often exhibit radioresistance through shared mechanisms [27–30]. Assessing chemotherapy IC50 in PDOs provides a consistent, high-throughput endpoint, and prior studies show close correlation between ex vivo chemo-resistance and poor clinical outcomes [24, 25]. This approach lays the groundwork for subsequent validation of promising methylation biomarkers against full TNT regimens in prospective cohorts. In this study, we pursue two main goals. First, we characterize the methylome in both RC tumor tissue and matched PDOs to investigate how epigenetic features correlate with ex vivo chemotherapy response. Second, we test the hypothesis that combining methylation profiles from both RC tumors and matched PDOs can significantly enhance predictive accuracy for chemoresistance. By demonstrating that methylation patterns shared between tumor and PDO inform treatment outcomes more reliably than either alone, we aim to establish a new framework for epigenetic biomarker discovery in rectal cancer. Ultimately, our findings could guide more personalized therapeutic strategies, improving the efficacy of total neoadjuvant therapy and reducing unnecessary toxicity for patients whose tumors are inherently resistant.

## Results

### Methylation consistency between tumors and matched PDOs

All 50 samples (18 primary tumor tissues and 32 organoids) passed our quality-control filters (see Methods and Supplementary Material for details) and were retained for methylation analysis. After removing probes on sex chromosomes, those with detection P-value > 0.01, and those overlapping known SNPs, a total of 771,964 CpG probes remained for downstream analyses.

We first assessed the reproducibility of methylation profiles among organoids derived from the same patient. In a correlation matrix (**Fig. 1A**), replicate organoid samples consistently exhibited high pairwise Spearman’s correlation coefficients, illustrating that organoids faithfully captured the patient-specific methylation landscape. Notably, correlations among three replicate organoids from a single tumor generally exceeded 0.96 (P < 0.001), underscoring the reliability of the organoid model.

**Figure 1.**
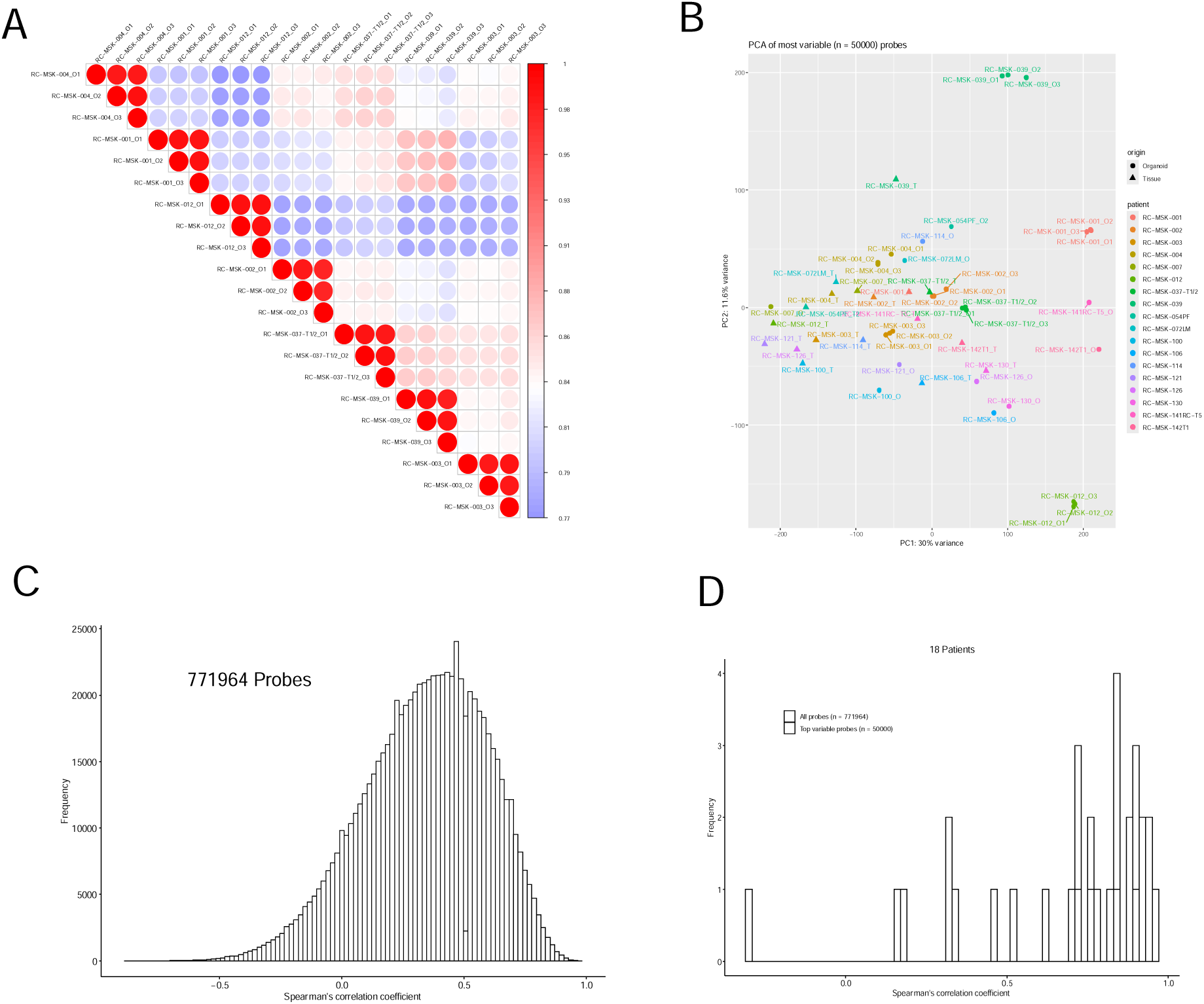
Methylation Profiling Consistency Between Rectal Cancer Tissues and Matched PDOs (A) Spearman correlation matrix showing high reproducibility among replicate organoid samples (n=7 patients with 3 replicates each). Darker red indicates stronger positive correlations. (B) Principal Component Analysis (PCA) of the top 50,000 most variable CpGs, demonstrating separate clustering of tumor (red) versus PDO (blue) samples along the first principal component (PC1), while replicate PDOs from the same patient cluster together along PC2. (C) Distribution of Spearman’s correlation coefficients across 771,964 CpG probes, highlighting in orange the subset exceeding a 0.50 correlation. This underscores broad agreement between tumor and PDO methylation patterns, particularly among highly variable CpGs. (D) Patient-specific correlation between each tumor sample and the mean profile of its matched PDO replicates. Most patients exhibit correlation coefficients >0.5, supporting the capacity of PDOs to recapitulate their parental tumor’s methylation landscape.

Next, we performed PCA on the top 50,000 most variable CpG probes from the 50 samples (**Fig. 1B**). Tissue- and organoid-derived samples formed two discrete clusters along the first principal component (PC1), suggesting a clear divergence in global methylation patterns between primary tumors and their matched PDOs. However, replicate organoids from the same patient clustered tightly together and displayed more variation along the second principal component (PC2)—highlighting both the unique epigenetic signature of the PDO cohort and the reproducibility of replicates derived from the same tumor. Given that each patient’s triplicate organoid set exhibited a highly reproducible methylation profile, the single replicate with the highest bisulfite conversion rate was selected for all subsequent analyses.

To better characterize the breadth of CpG site variability, we plotted the distribution of Spearman’s correlation coefficients for all 771,964 probes and then highlighted the subset of the top 227,101 probes with correlation coefficence greater than 0.5 (orange bars) (**Figure 1C**). The majority of these probes showed moderate-to-strong positive correlations between paired tissue and organoid samples. Notably, when restricting to the most variable sites, the correlation skewed higher, indicating that probes displaying large methylation differences across samples can still maintain consistent tumor–organoid patterns. This overall distribution reaffirms that rectal cancer PDOs capture both broad and high-variability methylation signals from their parental tumors.

To gauge how closely RC PDOs recapitulate the methylation patterns of the parental tumor, we compared each tumor sample’s profile to the mean methylation profile of its respective organoid replicates (**Fig. 1D**). Across all 771,964 CpG probes, the majority (∼97%) showed a positive Spearman’s correlation between tissue and PDO methylation intensities, indicating broad conservation of methylation states. When focusing on the top 50,000 variable CpG probes, the correlation strengthened further, consistently exceeding 0.50 for most patients (P < 0.05). Taken together, these results affirm that rectal cancer PDOs recapitulate the core methylation landscape of the parental tumors and underscore the utility of RC PDOs as a robust ex vivo model for investigating epigenetically mediated treatment responses.

### CpG selection for chemotherapy responsiveness

We investigated the relationship between CpG methylation (using M-values) and chemotherapy response (log-transformed FOLFOX IC50) by fitting linear models for each tissue type (tumor vs. PDO) individually, adjusting for age, sex, and batch (see Methods). We then extended the analysis to a joint linear mixed-effects model, incorporating patient-specific random effects to account for the correlation among multiple samples from the same individual. This allowed us to identify CpGs where methylation levels (M-values) showed a statistically significant association with log-transformed FOLFOX values across tissue and organoid samples. We derived a set of 745 top CpGs meeting FDR 0.05 threshold (**Supplementary Table 1**).

Figure 2 illustrates a heatmap of these 745 CpGs, with rows representing CpG sites and columns representing the 50 samples (18 tumor tissues and 32 organoids). The M-values range from −2 (blue) to +2 (yellow), highlighting the extent of hyper- or hypomethylation across the cohort.

**Figure 2.**
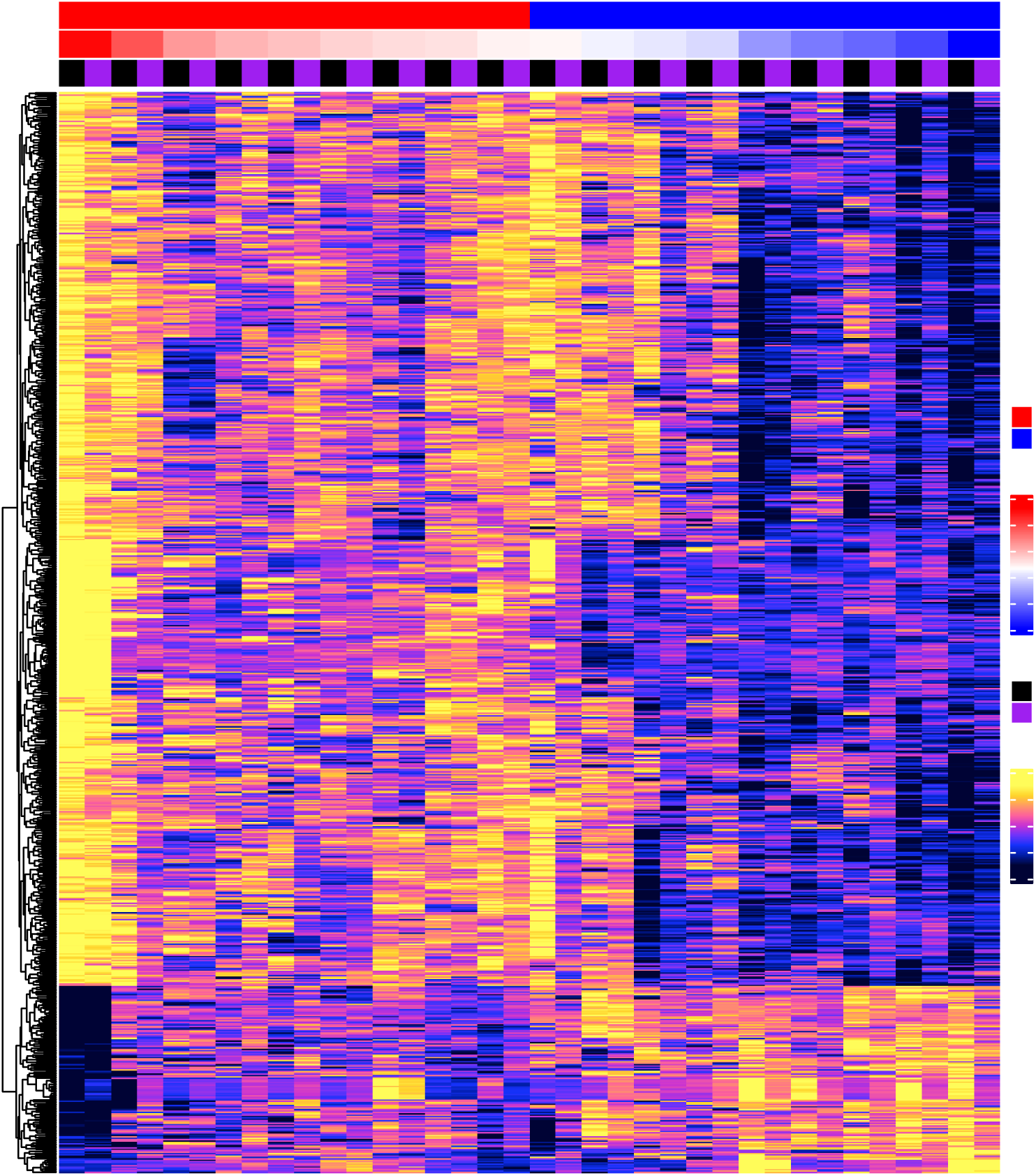
Heatmap of 745 CpGs Significantly Associated with FOLFOX IC50 Rows represent the 745 CpGs meeting the 5% FDR threshold in the joint linear mixed-effects model; columns include all 50 samples (18 tumor tissues and 32 PDOs). M-values range from −2 (blue, hypomethylation) to +2 (yellow, hypermethylation). The top annotation bar indicates each sample’s origin (tumor vs. PDO) and chemotherapy response level (high or low FOLFOX IC50). Hierarchical clustering shows distinct methylation patterns tied to chemo-resistance, with tumor and PDO samples interspersed but largely grouped by FOLFOX sensitivity.

Notably, tumor and PDO samples exhibit both shared and distinct patterns in these top CpGs, consistent with the finding from the joint model that many CpG sites display correlated methylation changes across sample types yet maintain tumor- or PDO-specific methylation signatures. The clustering dendrogram on the left indicates that resistant (high-IC50) and sensitive (low-IC50) samples often group into distinct subbranches, suggesting that these CpGs may underlie epigenetic differences contributing to variability in FOLFOX response.

**Table 1** highlights 14 CpGs that were significant (FDR <0.05) in the joint analysis but not significant in the tissue-only analysis (P__tissue_ > 0.05), which could not have been identified from the tissue samples alone. Interestingly, all 14 CpGs were hypermethylated with increasing FOLFOX IC50 values (i.e., higher chemoresistance). The majority of these CpGs (9 out of 14) were located in distal regions; only 5 CpGs were found in promoter regions. Among the genes associated with these 5 promoter-associated CpGs, *ZNF160*, *PTCHD3*, *ASTN1*, *PAX3*, and *HS3ST2*. *ZNF160* and *PAX3* are TFs that regulate genes involved in cell proliferation and apoptosis, influencing chemotherapy response[31–34]. *PTCHD3* is part of the Hedgehog signaling pathway, which is implicated in cancer progression and chemoresistance[35–37]. *ASTN1*, while known for neuronal migration, may affect cancer cell adhesion and migration, potentially altering chemotherapy efficacy[38]. *HS3ST2* modifies heparan sulfate proteoglycans, affecting growth factor signaling and interactions with the tumor microenvironment[39]. Given these findings, it is plausible to consider that hypermethylation of the promoters of these genes could lead to changes in expression, thus contributing to increased chemoresistance.

**Table 1.** Fourteen CpGs Identified Solely by the Joint Analysis but Not Tissue-Only List of CpGs with FDR <0.05 in the joint (tumor + PDO) linear mixed-effects model yet not significant (p >0.05) in the tissue-only analysis. For each CpG, columns include its genomic location, linear model estimates (coefficients, p-values, FDR), and annotation details (promoter vs. distal). Notably, all 14 CpGs are hypermethylated with higher FOLFOX IC50 (chemoresistance). Five CpGs map to promoter regions near *ZNF160*, *PTCHD3*, *ASTN1*, *PAX3*, and *HS3ST2*.

| CpG | Joint analysis of PDO and tissue samples |  |  |  | PDO-tissue sample corr. |  | chr | position | Annotations<br>GREAT_annotation |
| --- | --- | --- | --- | --- | --- | --- | --- | --- | --- |
|  | estimate | stdErr | pValue | fdr | estimate | pValue |  |  |  |
| cg08228914 | 0.694 | 0.165 | 2.55E-05 | 3.57E-02 | 0.358 | 1.45E-01 | chr19 | 53,103,284 | **ZNF160 (+149)** |
| cg04283864 | 0.716 | 0.174 | 4.00E-05 | 4.54E-02 | 0.521 | 2.84E-02 | chr6 | 4,188,951 | CDYL (-587483);ECI2 (-53356) |
| cg09544380 | 0.832 | 0.191 | 1.37E-05 | 2.64E-02 | 0.587 | 1.19E-02 | chr10 | 27,414,556 | **PTCHD3 (-190)** |
| cg25260543 | 0.867 | 0.210 | 3.74E-05 | 4.42E-02 | 0.647 | 4.61E-03 | chr7 | 101,303,510 | FIS1 (-58427);RABL5 (+18292) |
| cg14659404 | 1.251 | 0.288 | 1.42E-05 | 2.68E-02 | 0.657 | 3.84E-03 | chr1 | 177,164,882 | **ASTN1 (-194)** |
| cg02963248 | 1.407 | 0.300 | 2.82E-06 | 1.26E-02 | 0.662 | 3.57E-03 | chr2 | 66,582,647 | ETAA1 (-814671);MEIS1 (+147249) |
| cg14093610 | 1.029 | 0.251 | 4.05E-05 | 4.58E-02 | 0.670 | 3.06E-03 | chr2 | 222,297,946 | EPHA4 (-725657);**PAX3 (+1034)** |
| cg14670629 | 0.621 | 0.140 | 9.44E-06 | 2.30E-02 | 0.688 | 2.14E-03 | chr10 | 29,480,626 | LYZL1 (+191567);SVIL (+255174) |
| cg03877332 | 0.789 | 0.187 | 2.49E-05 | 3.51E-02 | 0.699 | 1.73E-03 | chr4 | 6,246,102 | JAKMIP1 (-45549);WFS1 (-23747) |
| cg09159223 | 0.613 | 0.149 | 3.75E-05 | 4.42E-02 | 0.759 | 4.02E-04 | chr7 | 35,979,077 | EEPD1 (-174070);SEPT7 (+178062) |
| cg00885682 | 1.000 | 0.245 | 4.55E-05 | 4.86E-02 | 0.771 | 2.76E-04 | chr16 | 22,814,427 | **HS3ST2 (+252)** |
| cg00737841 | 0.921 | 0.212 | 1.37E-05 | 2.64E-02 | 0.783 | 1.81E-04 | chr10 | 128,928,359 | MKI67 (-801976);MGMT (-538824) |
| cg05061450 | 0.906 | 0.220 | 3.88E-05 | 4.47E-02 | 0.800 | 9.34E-05 | chr10 | 123,355,457 | GPR26 (-310897);BUB3 (+201182) |
| cg13411311 | 0.909 | 0.220 | 3.67E-05 | 4.40E-02 | 0.806 | 6.95E-05 | chr2 | 240,156,571 | GPC1 (-279099);OTOS (-12011) |

### Differentially Methylated Region (DMR) analysis

To move beyond single–CpG comparisons and capture broader epigenetic shifts, we performed a region-based DMR analysis using the coMethDMR R package. Rather than examining each CpG site independently, this approach clusters adjacent CpGs (separated by ≤200 bp) into “contiguous genomic regions” and then identifies co-methylated subregions within these clusters. By summarizing the CpG M-values (logit-transformed β-values) as a median for each subregion, we tested their association with log-transformed FOLFOX IC50 values in a linear model—adjusting for potential confounders (age, sex, and batch). This strategy is advantageous for detecting functionally relevant epigenetic changes that span multiple CpGs, offering a more integrative view of how local methylation patterns may relate to chemotherapy response.

The list of 50 DMRs (**Supplementary Table 2**) highlights contiguous genomic regions where median methylation levels correlate significantly with FOLFOX response, even after adjusting for age, sex, and batch effects. Notably, many of these DMRs lie near transcription start sites (TSS), within active promoters (e.g., “Active TSS” or “Bivalent/Poised TSS”), or in potential regulatory elements such as enhancers or N/Shore regions. These annotations suggest that differential methylation in these regions could influence gene transcription by modulating chromatin accessibility.

Several DMRs are mapped to or near known cancer-related genes, including *CACNA1E*, *DIRAS1*, *TIGAR*, *CCND2*, and *SLC1A3*. In many instances, the positive estimates (for example, 0.7–1.6) indicate that higher methylation in these subregions is associated with greater resistance (i.e., higher log-FOLFOX IC50 values), whereas negative estimates imply a potential sensitizing effect when methylation is increased. The presence of multiple CpG sites within each region underscores how local “epigenetic blocks”—rather than single CpGs—may drive therapy outcomes.

Interestingly, several of the top DMRs overlap with shelf or shore contexts—areas flanking CpG islands—where methylation changes are often linked to subtle yet functionally significant regulation of nearby genes. Additionally, certain DMRs are annotated as “weak transcription” or “repressed polycomb” in their baseline chromatin states, implying that changes in methylation there might shift these regions toward (or away from) an active regulatory environment.

Overall, these region-level findings reinforce the notion that epigenetic regulation near key regulatory elements could modulate the tumor’s response to chemotherapy. By zooming in on contiguous CpGs in a biologically coherent way, the DMR approach complements the single-CpG analyses and uncovers broader methylation patterns potentially critical to FOLFOX sensitivity or resistance in rectal cancer.

### Pathway and Geneset Analysis

We applied a gene set enrichment analysis (GSEA) using *methylRRA* in the methylGSA R package to explore the biological pathways enriched with CpG methylation changes linked to FOLFOX response. After aggregating bacon-corrected *p*-values at the gene level, we ran a pre-ranked GSEA on the KEGG and Reactome databases, considering pathways with a false discovery rate (FDR) below 25% (**Supplementary Table 3)**. In KEGG, top-ranking pathways included *Focal adhesion* and *ECM–receptor interaction*, both pointing to the involvement of ECM- and integrin-related genes (*COL1A1*, *ITGA4*, and *FN1*, among others) in potentially modulating tumor cell sensitivity or resistance to FOLFOX [11, 40–45]. The *Calcium signaling pathway* also stood out, driven by a range of voltage-gated channels and G protein–coupled receptors (*CACNA1E*, *RYR*, *ADRA1*), suggesting that epigenetic shifts in calcium homeostasis might affect proliferation and apoptosis under drug stress [46]. Notably, *One carbon pool by folate* emerged as particularly relevant to 5-FU–based regimens that target the folate cycle, implicating genes like *ALDH1L1* and *MTHFD1L* in shaping FOLFOX efficacy [47]. Other enriched KEGG sets, including *proximal tubule bicarbonate reclamation* and *chronic myeloid leukemia*, reinforce the multifactorial nature of chemotherapy response, implicating both systemic and cancer-specific epigenetic regulation.

In the Reactome database, pathways related to developmental and signaling processes featured prominently. *Activation of anterior HOX genes* and *Activation of HOX genes during differentiation* highlight the potential significance of epigenetic regulation within the HOX cluster (*HOXA3*, *HOXB2*), which can influence cell fate and proliferation [48, 49]. Epigenetic changes involved in *ERK/MAPK targets* and *RSK activation* likewise point to the possibility that MAPK–RSK cascades mediate tumor cell survival under chemotherapeutic pressure [50]. Additionally, *p75 NTR receptor-mediated signaling* and *Integrin cell surface interactions* both suggest that reconfiguration of extracellular communication—via adhesion molecules, senescence signals, and apoptotic modulators—could underlie variations in FOLFOX responsiveness [51]. Collectively, these findings reveal that Folfox-associated methylation changes are not confined to a single pathway but span diverse processes, including ECM remodeling, HOX gene governance, and cell survival strategies, underscoring how epigenetic plasticity across multiple networks might shape rectal cancer chemosensitivity.

### Survival prediction based on DNAm

Fig. 3 shows the Kaplan-Meier curves for patients in the highest and lowest quartiles of methylation risk scores. While the methylation risk scores based on rectal cancer PDO-only or tissue-only data did not predict progression-free survival (Fig. 3A**,B**), scores derived from the joint analysis of both tissue types (Fig. 3C) reached marginal significance (p=0.05). Notably, the methylation risk scores based on the 14 CpGs that were significant in the joint analysis but not in the tissue-only analysis (Fig. 3D) showed the strongest predictive accuracy for progression-free survival (p=0.019).

**Figure 3.**
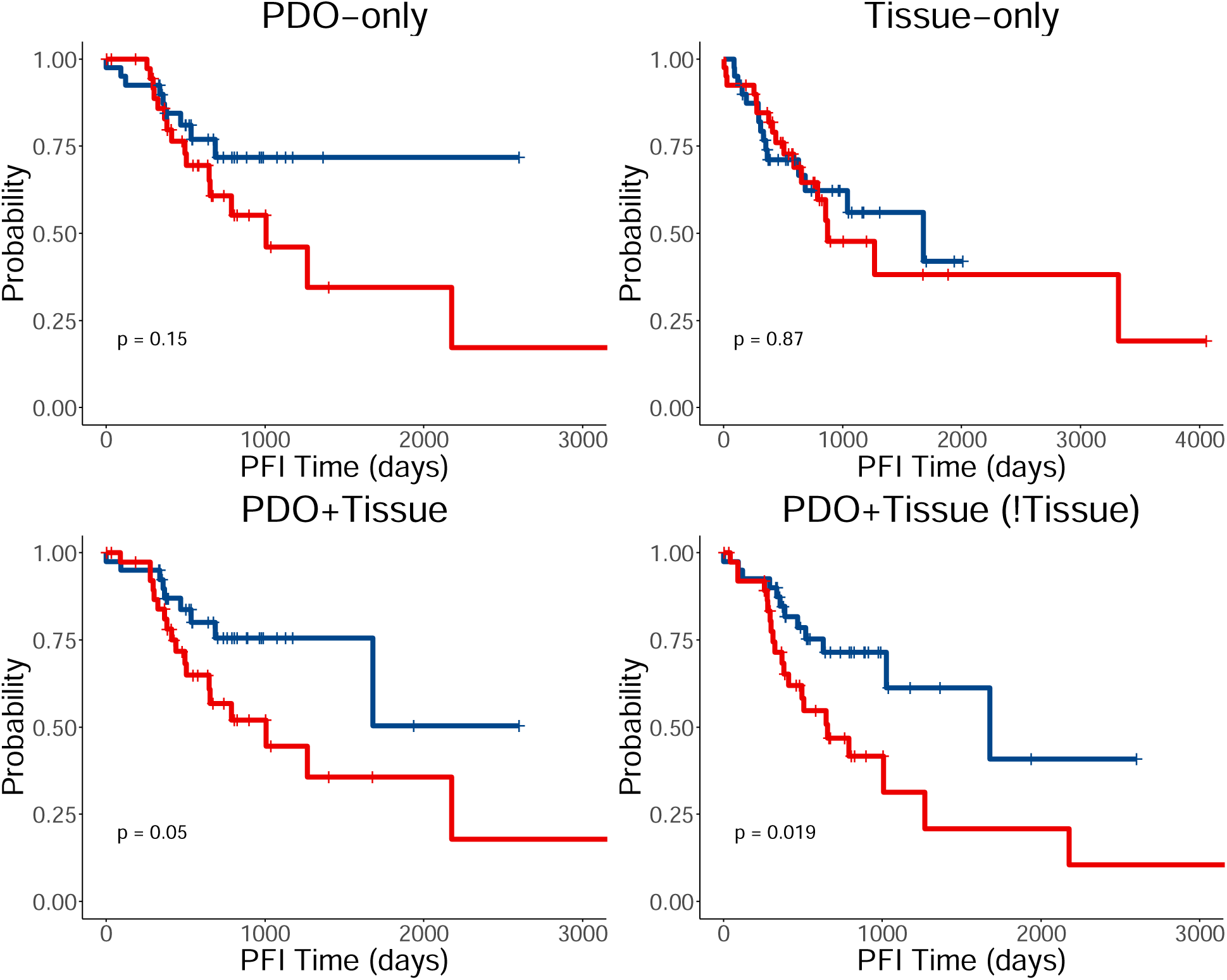
Survival Analyses Using Methylation Risk Scores (MRS) in an Independent RC Cohort Kaplan-Meier curves evaluating progression-free survival (PFI) for patients with high (blue) vs. low (red) MRS, stratified by quartiles. (**A**, **B**) MRS derived from PDO-only or tissue-only CpGs shows no significant PFI difference. (C) MRS from the joint tissue–PDO analysis yields marginal significance (p=0.05). (D) MRS based on 14 “joint-only” CpGs demonstrates the strongest PFI discrimination (p=0.019), indicating the added predictive value of integrating tumor and PDO epigenetic data.

In summary, integrating DNAm data from both tumor and matched PDOs yields more robust predictive signatures for chemotherapy response (progression-free survival) than analyzing either tissue type alone. This integrated approach can highlight critical CpGs and gene targets otherwise missed by single-sample analyses, strengthening our rationale for combining rectal cancer PDO and tumor data to identify DNAm biomarkers of TNT response.

## Methods

### Human RC tissue and patient-derived organoid

A total of 18 patients diagnosed with rectal cancer (RC) at Memorial Sloan Kettering Cancer Center were selected in our study. For each patient, one tumor tissue was obtained before treatment, and at least one matched patient-derived organoid (PDO) samples were generated. In specific, for each of the eleven out of 18 patients, one single PDO was generated, while to validate the replicability of PDO model, each of the remaining seven patients had triplicated organoid samples. Endoscopic tumor biopsies obtained with biopsy forceps were obtained from RC patients undergoing standard of care diagnostic biopsy or on-treatment biopsy in the outpatient setting. The specimens are collected per protocol and tissue was prioritized as follows: sample sent to pathology, sample for PDO, and remaining saved for banking. Tissues are chopped into ∼1-mm pieces in cold PBS + antibiotics, and after tissue processing, tumor cells are resuspended in 300 μL of Matrigel and plated onto 24-well culture plates (40 μL droplet per well). After the gel solidifies, 700-800 μL of culture medium is added to wells and incubated at 37°C. PDOs are established and maintained in Matrigel as described in Ganesh et al.[24]. Consequently, 18 tissues and 32 matched organoids were involved in this study and were then RNA-sequenced (see below).

### Preparation and methylation array of tissue and organoid samples

Surgically resected rectal tissue or biopsy tissue was washed with ice-cold PBS-Abs buffer (phosphate-buffered saline with antibiotic-antimycotic (Gibco™), gentamicin (Gibco™) and plasmocin (Invivogen). The tissue was divided into two parts. One part was flash-frozen in liquid nitrogen and then preserved at −80°C for later purposes. The other part was processed for tumoroid culturing purposes, as reported[22]. The patient-derived tumoroids were cultured and expanded in Matrigel for three passages. To extract DNA, well-growing tumoroids in 4-6 Matrigel domes (40ul Matrigel/dome) were dissociated with TrypLE Express (Gibco™) as reported[22]. The obtained pellet was further washed with 5 ml of ice-cold PBS. Then, DNA was extracted from the tumoroid pellet using the AllPrep DNA kit (Qiagen) following the manufacturer’s instructions. If the DNA needed to be extracted separately, then the DNeasy Blood & Tissue Kit (Qiagen) was used. The tissue DNA was extracted from the flash-frozen specimen using the AllPrep DNA ki..

Samples were bisulfite converted according to Illumina’s specifications. DNA input for Bisulfite conversion was 500ng. DNA concentration was assessed via Qubit and quality assessed via Tape station 4200. The customer’s samples were processed using the MethylationEPIC_v-1-0 bead chip from Illumina with 866,895 CpGs.

### Pre-processing of DNA methylation data

The samples were measured using the Infinium MethylationEPIC BeadChip, and **Supplementary Table 4** shows the number of CpGs and samples removed at each quality control step. Quality control for CpG probes included removing probes with detection *P-*value > 0.01 in any sample using the minfi R package, removing probes that are cross-reactive [52], having a single nucleotide polymorphism (SNP) with minor allele frequency (MAF) ≥ 0.01 present in the last 5 base pairs of the probe, or located on the sex chromosomes using the DMRcate R package (with function rmSNPandCH and parameters dist = 5, mafcut = 0.01). For quality control of samples, we removed samples with low bisulfite conversion efficiency (i.e., < 85%). We also removed 1 sample outlier with an extremely high folfox level detected by the PCA plot. Given the high reproducibility of organoid methylation levels, when multiple organoid samples were available for a patient, we selected the organoid sample with the highest bisulfite conversion rate. The quality-controlled methylation datasets were then subjected to the β-mixture quantile normalization (BMIQ) procedure[53], as implemented in the wateRmelon R package[54], which normalizes beta values of type 1 and type 2 design probes within the Illumina arrays.

### Statistical analysis of individual CpGs

The association between CpG methylation levels and log-transformed Folfox values was assessed using a linear statistical model for each tissue type (tissue or tumoroid) separately. Given that methylation M-values (logit transformation of methylation beta values) have better statistical properties (i.e., homoscedasticity) for linear regression models[55], we used the M-values as the outcome variable. This model also included log-transformed Folfox values as the main independent variable and age, sex, and batch as covariates. We used the limma R package, which uses the empirical Bayes method and is suitable for analyzing small sample size studies.

The joint analysis of both tissue and tumoroid samples was analyzed using a linear mixed-effects model. This model included the same fixed effects (log_Folfox, sex, age, and batch) but also included patient random effects to account for the correlations arising from multiple samples from the same subject. The dupcor() function in the limma R package was used to account for within-sample correlations.

### Region-based analysis

To identify differentially methylated regions significantly associated with age at death, we used the coMethDMR R package. The “contiguous genomic regions” are genomic regions on the Illumina array covered with clusters of contiguous CpGs where the maximum separation between any two consecutive probes is 200 base pairs. First, coMethDMR selects co-methylated sub-regions within the contiguous genomic regions. Next, we summarized methylation M values within these co-methylated sub-regions using medians and tested them against log-transformed Folfox values, while adjusting for age, sex, and batch as covariate variables.

### Inflation and multiple testing correction

The *bacon* method was also used to obtain inflation-corrected effect sizes, standard errors, and *P-*values for each analysis.[56] To account for multiple comparisons, False Discovery Rate (FDR) based on the method of Benjamin and Hochberg [57] were computed to adjust bacon-corrected *P-*values. We considered individual CpGs with 5% FDR to be statistically significant. For DMRs, we additionally required significant DMRs to have at least 3 CpGs.

### Functional annotations and enrichment analyses

The significant DNAm differences at individual CpGs and DMRs were annotated using the Illumina annotations and the GREAT (Genomic Regions Enrichment of Annotations Tool) software[58] with default parameters. To identify biological pathways enriched with Folfox-associated DNA methylation, we used the methylRRA function in the methylGSA R package[59] (version 1.14.0). The pathway analyses were performed separately for each of the three analyses (tissue only, tumoroid only, and joint analysis). In each analysis, we used the bacon-corrected *P*-values from individual CpGs analysis described above as the input for methylGSA. Briefly, methylGSA first computes a gene-wise ρ value by aggregating *P-*values from multiple CpGs mapped to each gene. Next, the different number of CpGs on each gene is adjusted by Bonferroni correction. Finally, a Gene Set Enrichment Analysis[60] (in pre-rank analysis mode) is performed to identify pathways enriched with significant Folfox-associated DNAm. We analyzed pathways in the KEGG[61] and REACTOME[62] databases. Because of the relatively smaller number of gene sets being tested, a 25% FDR significance threshold, instead of the conventional 5% FDR, was suggested to be the default significance threshold for GSEA (https://software.broadinstitute.org/cancer/software/gsea/wiki/index.php/FAQ). Therefore, we considered pathways with FDR < 0.25 as statistically significant.

### Survival analysis

We further evaluated the utility of DNAm in predicting progression-free survival (PFI) in subjects who underwent chemotherapy, using data from the TCGA dataset (n = 160 colorectal cancer patients). Since direct chemosensitivity data was unavailable in the TCGA dataset, we used PFI status as a surrogate marker for chemotherapy success. To this end, we computed methylation risk scores (MRS) by summing the methylation M-values of CpGs that were significant (FDR < 0.05) in our pilot analysis, weighted by their estimated effect sizes for log-transformed Folfox values in the linear models described above.

## Discussion

Our findings underscore the pivotal role of epigenetic modifications in shaping chemotherapy response in rectal cancer, particularly when leveraging the complementary strengths of both tissue and patient-derived organoid (PDO) samples. Prior studies have established the importance of DNA methylation—ranging from locus-specific changes in genes like *MLH1*, *MGMT*, or *TFAP2E* to broader methylation phenotypes such as CIMP—in colorectal tumor biology and therapy outcomes. However, many methylation biomarkers exhibit inconsistent validation, and few have translated into routine clinical practice. By integrating matched tumor– PDO data, we addressed a core challenge in epigenetic biomarker discovery: separating genuine therapy-relevant signals from confounding artifacts. Our results show that methylation patterns associated with FOLFOX response can be more robustly identified when the same epigenetic signal recurs in both a patient’s tumor and its corresponding organoid. This joint approach uncovered specific CpGs and differentially methylated regions (DMRs) that would have been missed by tissue-only analysis, offering new insights into potential drivers of chemoresistance.

In particular, we observed that a subset of CpGs—hypermethylated in correlation with FOLFOX IC50—involves genes and pathways not previously implicated in neoadjuvant rectal therapy. Notably, these included distal CpG sites, providing a reminder that epigenetic control often extends beyond classical promoter regions to enhancers, shores, and other regulatory elements. The DMR analysis further reinforced this point, revealing contiguous genomic blocks, sometimes spanning multiple CpGs, linked to drug response. Such region-level shifts in methylation can lead to subtle but coordinated changes in gene regulation, possibly affecting entire biological modules rather than a single gene promoter. Supporting this, our pathway enrichment analyses identified recurrent themes—ECM–receptor interaction, focal adhesion, and calcium signaling among them—indicating that epigenetic disruption of tumor cell– microenvironment communication and intracellular homeostasis may govern response to cytotoxic drugs like 5-FU and oxaliplatin. Interestingly, we also found that folate-related and HOX regulatory pathways were enriched, mirroring earlier reports that folate metabolism heavily influences 5-FU efficacy and that HOX gene dysregulation can direct cell fate decisions in therapy settings.

Crucially, beyond simply cataloging CpGs and pathways, we tested their clinical relevance by constructing methylation risk scores (MRS) and evaluating their ability to predict progression-free survival in an independent patient cohort. While single-sample–based MRS (from either tumor or PDO alone) lacked significant prognostic value, the MRS arising from our joint model achieved marginal significance, and the subset of 14 “joint-only” CpGs demonstrated the strongest association with patient outcomes. This highlights the practical benefit of simultaneously analyzing ex vivo PDO data and in vivo tumor tissue—particularly for capturing epigenetic determinants of chemoresistance. Still, some limitations must be acknowledged. First, our sample size, though in line with other PDO-based methylation studies, remains modest; larger cohorts are needed to refine these biomarkers for clinical use. Second, we did not account for intratumoral heterogeneity that might persist even within PDO cultures, nor did we incorporate microenvironmental factors beyond the epithelial compartment. Future efforts could integrate immune components or fibroblasts in organoid co-cultures to capture additional dimensions of therapy response.

Finally, while our approach focuses primarily on FOLFOX-based chemoresponse, validating these methylation biomarkers in the full TNT setting—including radiotherapy—will be crucial for broader clinical applicability in rectal cancer. Overall, this work establishes a proof-of-concept that combining DNA methylation profiles from both tumor tissue and matched PDOs can yield more robust biomarkers of rectal cancer chemoresistance. By bridging preclinical models with patient-derived samples, we can identify epigenetic changes genuinely related to therapy outcomes and thus reduce false positives that often plague single-sample omics studies. If validated in larger, prospective cohorts, these findings may lay the groundwork for epigenetically guided stratification of rectal cancer patients, sparing non-responders from ineffective treatments while personalizing regimens for those most likely to benefit.

## Data Availability

All data produced in the present study are available upon reasonable request to the authors

https://portal.gdc.cancer.gov/

## DECLARATIONS

### Availability of data and materials

The datasets analyzed in this study are available upon the request to the corresponding authors

